# Physical activity buffers physiological stress during high emotional distress: a wearable-derived prospective cohort study

**DOI:** 10.64898/2026.04.05.26350215

**Authors:** Cadence Pinkerton, Yuqing Guo, Annie Qu

## Abstract

**Background:** Digital phenotyping using wearable devices and ecological momentary assessment (EMA) enables continuous, real-world monitoring of physiological and emotional states, but identifying high-risk stress states in real time remains challenging. We examined day-level associations between emotional distress and heart rate variability (HRV), and assessed whether daily physical activity modifies this relationship using longitudinal wearable and EMA data.

**Methods:** The Smart Momentary Interactive Longitudinal Evaluation Study (SMILES) was a prospective cohort study conducted among STEM graduate students in the U.S. in 2025. Participants wore an Oura Ring Generation 3 continuously for five months and completed daily EMA surveys assessing emotional distress. The primary outcome was nightly HRV measured as the root mean square of successive differences and log-transformed for analysis. Quantile regression within a quadratic inference function framework was used to estimate associations at the 25th, 50th, and 75th percentiles of HRV, accounting for within-participant correlation and time-varying covariates.

**Findings:** Thirty-one participants contributed 1,724 person-days of observation. High emotional distress was associated with lower HRV across the HRV distribution, with the strongest association observed at the lower HRV quantile (*β* = -0·094, 95% CI: [-0·111, -0·078]). A significant interaction between daily step count and emotional distress was observed across quantiles, such that higher physical activity was associated with higher HRV on high emotional distress days but not on low-to-moderate distress days.

**Interpretation:** Integration of wearable-derived physiological data with EMA enables real-time identification of high-risk stress states in naturalistic settings. The observed buffering effect of physical activity during periods of elevated emotional distress suggests that wearable-guided, personalized just-in-time adaptive interventions, such as physical activity prompts, could be deployed to improve autonomic regulation and mental health.

**Funding:** National Cancer Institute (R01: CA297869) and National Science Foundation (DMS: 2515275).

Research in context

Evidence before this study
We searched PubMed, Google Scholar, and Web of Science for studies published up to March 1, 2026, using combinations of the terms “wearable devices,” “heart rate variability,” “stress,” “physical activity,” “ecological momentary assessment,” “digital phenotyping,” and “graduate students.” Previous studies have shown that graduate students experience high levels of psychological distress and are at increased risk of anxiety, depression, and burnout. Heart rate variability (HRV) is a well-established biomarker of autonomic regulation and physiological stress, and physical activity has been associated with improved HRV and stress resilience. Wearable devices and ecological momentary assessment (EMA) have enabled real-time measurement of physiological and emotional states in naturalistic settings, and several studies have used wearable devices to examine stress and physical activity in daily life. However, most existing studies rely on cross-sectional or mean-based analyses and do not examine whether the association between emotional distress and physiological stress varies across physiological states or over time within individuals. In addition, few longitudinal studies have integrated wearable-derived physiological data with repeated EMA to examine day-level associations between emotional distress, physical activity, and physiological stress in real-world academic settings.

Added value of this study
Together with existing literature, our findings suggest that wearable devices combined with ecological momentary assessment can be used to monitor physiological stress in real time and identify periods of elevated stress risk. The observation that physical activity is associated with improved physiological regulation specifically on high emotional distress days suggests that wearable-based monitoring systems could support personalized, just-in-time adaptive interventions that prompt physical activity during periods of elevated stress. Such approaches may help reduce chronic stress and improve mental health in graduate students and other high-risk populations. More broadly, this study demonstrates the potential of digital phenotyping approaches that integrate wearable sensors and self-reported data to inform personalized and adaptive mental health interventions in real-world settings.

Implications of all the available evidence
Together with the existing literature, our findings indicate that wearable technologies combined with ecological momentary assessments can help identify daily stress in graduate students as it occurs. The observation that physical activity may act as a buffer between emotional distress and physiological stress can help inform adaptive, just-in-time digital health interventions, reducing the risk of burnout, anxiety, and depression among STEM graduate students. Universities and student health programs may consider student well-being initiatives that leverage wearable devices to enable individualized and timely intervention. Future studies could evaluate whether a physical activity prompt delivered following a high emotional distress day can improve autonomic regulation and psychological well-being in graduate students.

## Introduction

Graduate students experience disproportionately high levels of psychological stress, with documented associations with anxiety, depression, and emotional distress [1]. Evidence suggests that individuals pursuing higher education report more symptoms of mental disorders than those who do not [2]. In particular, graduate students exhibit moderate to severe depressive and anxiety symptoms at rates exceeding those observed in the general population, with an estimated 20–50% reporting clinically relevant symptoms [3]. Persistent exposure to stress may contribute to burnout, characterized by emotional exhaustion, cynicism, and reduced professional efficacy [4].

Stressors in graduate education are multifaceted and include competitive academic environments, performance pressure, financial strain, and work–life imbalance. These stressors are not evenly distributed. Underrepresented minority (URM) students may encounter differential treatment and diminished sense of belonging, contributing to heightened imposter syndrome and stress exposure [5]. First-generation and international students face additional academic, cultural, and structural challenges that may compound existing stressors [6, 7, 8, 9]. Despite growing recognition of these disparities, much of the evidence relies on cross-sectional or retrospective self-report measures.

Real-time identification of high-risk stress states remains limited. Although validated self-report instruments such as the Perceived Stress Scale are widely used, subjective measures are susceptible to recall bias and reporting error [10]. Chronic stress may impair working memory and cognitive processing, potentially limiting the reliability of retrospective stress assessments [11]. As stress encompasses both emotional and physiological dimensions, integrating objective biological measures with real-time self-reported data may provide a more complete understanding of stress dynamics in naturalistic settings.

Heart rate variability (HRV) reflects beat-to-beat fluctuations in cardiac intervals and autonomic nervous system regulation [12]. The root mean square of successive differences (RMSSD) is a commonly used metric of parasympathetic activity and vagal tone [13]. Higher HRV is associated with greater adaptive capacity and stress resilience, whereas lower HRV is linked to chronic stress and impaired autonomic flexibility [14]. Emotional distress is consistently associated with reduced HRV, reflecting heightened sympathetic activity and diminished vagal regulation [14, 15, 16]. In contrast, physical activity is positively associated with HRV through enhanced parasympathetic tone and autonomic balance [17, 18]. Age also contributes to HRV variation, with younger individuals generally exhibiting higher HRV and greater autonomic flexibility [19]. These relationships support HRV as an integrative biomarker of emotional and behavioral processes.

Advances in wearable technologies enable continuous, non-invasive monitoring of physiological signals in real-world settings. Consumer-grade devices such as the Oura Ring enable longitudinal measurement of sleep, physical activity, and cardiovascular function outside clinical environments [20]. These technologies support digital phenotyping approaches, in which passively collected physiological data and ecological momentary assessments (EMA) can be used to characterize dynamic health states. EMA enables repeated sampling of emotional states in real time and in natural environments [21], and captures short-term fluctuations relevant for stress and well-being [22]. However, existing approaches do not enable identification of high-risk stress states in real time or inform adaptive, personalized interventions.

The Smart Momentary Interactive Longitudinal Evaluation Study (SMILES) integrates longitudinal wearable-derived physiological data with daily self-reported emotional assessments to investigate the relationship between emotional distress and autonomic regulation among STEM graduate students. Beyond estimating average associations, we examine heterogeneity across the distribution of HRV to determine whether these relationships differ across physiological states. We further assess whether daily physical activity moderates this relationship, with the goal of informing real-time, personalized intervention strategies.

We hypothesized that higher emotional distress would be associated with lower nightly HRV, that this association would be strongest on days with low HRV, and that greater daily physical activity would attenuate this association.

## Methods

### Study design and participants

The SMILES study was designed as a prospective longitudinal cohort study. Ethical approval was obtained from the University Institutional Review Board prior to participant recruitment (IRB number: 5077). STEM graduate students at a public University in the U.S. were recruited using convenience sampling through IRB-approved flyers posted across campus and graduate housing facilities. Additionally, IRB-approved recruitment emails were distributed to faculty members in STEM departments for dissemination to graduate students.

Eligible participants were aged 18 years or older, currently enrolled as graduate students or postdoctoral scholars in STEM disciplines at this public University in the U.S., owned a smartphone with Wi-Fi access, and were willing to wear a Generation 3 Oura Ring wearable device over a five-month study period. Students were excluded if they reported medical conditions requiring ongoing treatment or intervention that could substantially affect sleep quality or physiological stress measures, or if they were pregnant at the time of enrollment.

Eligibility screening was conducted through an interest survey administered via REDCap (a secure data collection platform). Eligible individuals received an IRB-approved Study Information Sheet and an informed consent document. After verbal informed consent was obtained, participants met with a research coordinator (RC) to receive their Oura Ring Gen3 and instructions for device use.

A total of 37 students met eligibility criteria and were enrolled in the study. Three participants withdrew for personal reasons, resulting in 34 participants who provided wearable-derived physiological data, including sleep, physical activity, and heart rate variability. Of these, 31 participants completed both the EMA and provided wearable-derived physiological data, comprising the final analytic cohort.

The final cohort included 31 graduate trainees (25 PhD students, four master’s students, and two postdoctoral scholars). Participants self-identified as Asian (51·6%), White (38·7%), and Hispanic/Latino or Black/African American (9·7%).

### Procedures

At baseline, demographic information was collected via REDCap, including age, sex assigned at birth (male or female), education level, and race/ethnicity. Race and ethnicity were self-reported using categories adapted from the US Census Bureau (Black or African American; Hispanic or Latin/o/a/x; American Indian or Alaska Native; White; Native Hawaiian or Other Pacific Islander; South Asian; East Asian; Southeast Asian; Middle Eastern or Arab; mixed race; other; unknown; or decline to answer), with participants permitted to select all that applied.

The Generation 3 Oura Ring (Oura Health Oy, Oulu, Finland) is a waterproof, multi-sensor wearable device that continuously measures sleep, heart rate variability, and physical activity [23]. The device uses green, red, and infrared light-emitting diodes to estimate heart rate variability, while a three-dimensional accelerometer captures movement, daily activity, and step counts. Data are synchronized via Bluetooth to the Oura mobile application. During enrollment, participants installed the application under research coordinator (RC) supervision, selected ring size, and received standardized instructions for device use and synchronization.

Participants were instructed to remove the ring only when necessary (e.g., charging). The device collected nocturnal physiological measures, including the primary outcome, average nightly heart rate variability (HRV), quantified using the root mean square of successive differences (RMSSD). HRV was selected a priori as a marker of autonomic function and physiological stress. Additional measures included lowest nighttime heart rate, average breathing rate, body temperature deviation, recovery index, and time spent in sleep stages (deep, rapid eye movement [REM], and light sleep).

Body temperature and recovery index are reported on a 1–100 scale. The body temperature score reflects deviations from an individual’s baseline nocturnal temperature [24], and the recovery index reflects the timing of the lowest heart rate during sleep relative to total sleep duration, with higher scores indicating earlier cardiovascular recovery [25].

Participants also downloaded the Qulab EMA mobile application, developed by the study team and available for iOS and Android devices. The application collected ecological momentary assessment (EMA) responses and additional baseline characteristics, including international student and first-generation student status. Participants completed a daily EMA assessing emotional states using a validated 12-item inventory comprising positive (healthy, content, cheerful, excited, safe) and negative (tired, overwhelmed, lonely, angry, sad, worried, stressed) emotions [26].

Each day, participants received a reminder at a self-selected time. Items were presented as, “How [emotion] do you feel right now?” and rated on a 5-point Likert scale from 0 (“Not at all”) to 4 (“A lot”). Responses were categorized as high versus low-to-moderate emotional distress. Covariates included age, sex, international student status, first-generation student status, average breathing rate, lowest heart rate, body temperature, recovery index, and sleep stage durations (deep and REM).

## Statistical Analysis

The primary outcome was average nightly heart rate variability (HRV). The intensive longitudinal design yields 1,724 observations nested within 31 individuals, enabling within-person inference over time and reducing between-person confounding, an approach increasingly used in digital health research to understand dynamic health processes in real-world settings. Because HRV distributions are typically right-skewed and heterogeneous, we used quantile regression [27] to estimate covariate effects across the conditional HRV distribution. Results are presented for the 25th (*τ* = 0.25), 50th (*τ* = 0.5), and 75th (*τ* = 0.75) percentiles.

Because repeated HRV measurements within individuals are correlated, we used the Quadratic Inference Function (QIF) framework [28, 29] to obtain consistent and asymptotically efficient estimators without specifying a full parametric mixed-effects model. Compared with generalized estimating equations [30], QIF avoids estimating nuisance correlation parameters and improves efficiency under misspecification.

For subject *i* = 1, …, *n* observed at longitudinal time points *j* = 1, …, *m*_*i*_, let *Y*_*ij*_ denote HRV and *X*_*ij*_ ∈ ℝ^*p*^ the covariate vector. For a given quantile level *τ* ∈ (0, 1), we assumed the conditional *τ* th quantile

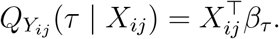

Let *r*_*i*_(*β*_*τ*_) = *y*_*i*_ − *X*_*i*_*β*_*τ*_ be the residual vector for subject *i*. Because the quantile score function is non-differentiable, we applied an induced smoothing function to enable stable optimization. The subgradient was replaced with

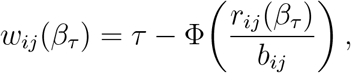

where Φ(·) is the standard normal cumulative density function (CDF) and 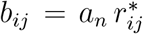 is an observation-specific bandwidth with *a*_*n*_ = *n*^−1*/*2^ and 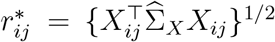 [31]. The covariate moment matrix 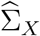 was estimated using the empirical second moment of all design rows pooled across subjects, with a small ridge term added for numerical stability.

To accommodate irregularly spaced multivariate observations common in mobile health (mHealth) data, we specified a continuous-time autoregressive correlation structure of order 1 [30]. This structure is more appropriate than discrete AR(1) assumptions when observation intervals vary due to missing responses or the nature of multi-resolution wearable data [32]. For observation times *t*_*ij*_, correlation was modelled as Corr(*Y*_*ij*_, *Y*_*ik*_) = exp{−*ϕ*|*t*_*ij*_ − *t*_*ik*_|} with fixed decay parameter *ϕ >* 0. The QIF extended score for each subject was constructed using *K* = 3 basis matrices 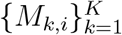 where 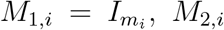, *M*_2,*i*_ with off-diagonal entries exp{−*ϕ*|*t*_*ij*_ − *t*_*ik*_|} and zeros on the diagonal, and a local adjacency matrix *M*_3,*i*_ indicating whether |*t*_*ij*_ − *t*_*ik*_| is within a data-driven neighborhood width, with zeros on the diagonal. These basis matrices provide a flexible approximation to the inverse working correlation matrix within the QIF framework [28, 29].

The stacked estimating equations were

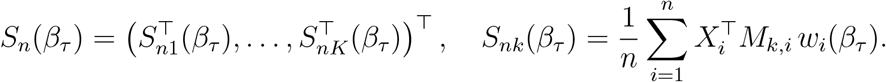

where 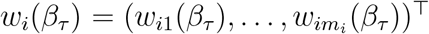 denotes the vector of smoothed quantile scores for subject *i*.

The QIF objective was

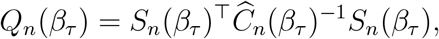

where 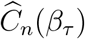 is the empirical covariance of the stacked subject-level score functions, with ridge regularization. Parameters *β*_*τ*_ were estimated by minimizing *Q*_*n*_(*β*_*τ*_) using a BFGS quasi-Newton algorithm, initialized by the pooled independent quantile regression estimate.

The variance of the regression coefficients was estimated using an asymptotic sandwich estimator based on the numerical Jacobian 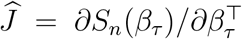 evaluated at 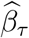 and the estimated covariance 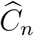:

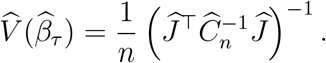

Standard errors were obtained from the diagonal of 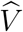, and two-sided Wald p-values were computed using the standard normal distribution based on the Wald test statistic 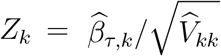 for each regression coefficient. Models were fitted separately for *τ* = 0.25, 0.50, and 0.75 to assess distributional heterogeneity, with *α* = 0.05.

A daily emotional distress score was derived from the 12-item EMA inventory. Positive items were reverse-coded and summed with negative items to yield a composite score ranging from 0 to 48, with higher values indicating greater distress. For primary analyses, emotional distress was categorized as high (≥ 21) versus low–moderate (*<* 21), with the threshold defined as the 75th percentile of the emotional distress distribution. Daily step count was included as a continuous measure of physical activity.

The quantile regression model was adjusted for demographic characteristics and nightly physiological covariates, including body temperature score and recovery index:

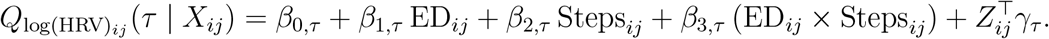

The interaction term was included to assess whether the association between physical activity and HRV differed by emotional distress level, where *Z*_*ij*_ denotes the vector of adjustment covariates including age, sex, international student status, first-generation student status, average breathing rate, lowest heart rate, body temperature, recovery index, deep sleep duration, and REM sleep duration.

Days with missing emotional distress scores or Oura-derived physiological measures were excluded. Two observations with step counts exceeding 30,000 steps were excluded as outliers. Additional analyses were conducted using different quantiles (*τ* = 0.1, 0.9) or modeling emotional distress as a continuous variable. Results from these analyses were consistent with the primary findings across both lower and upper quantiles. Similar trends were observed when emotional distress was modeled as a continuous variable rather than categorized. For interpretability, we present results based on the categorical emotional distress variable and the primary quantiles (*τ* = 0.25, 0.5, and 0.75).

All analyses were performed in R version 4.5.2.

## Results

The final sample included 31 participants contributing 1,724 person-days of observation. Participants ranged in age from 23 to 42 years (mean 28·10, SD 4·67), and 32·3% were male. Approximately 41·9% were international students and 35·5% identified as first-generation college students. Participant demographic characteristics are summarized in Table 1.

**Table 1:** Participant Characteristics.

| Characteristic | Overall (N = 31) |
| --- | --- |
| Age, mean (SD) | 28.10 (4.67) |
| Male, n (%) | 10 (32.3) |
| Racial background, n (%) |  |
| Asian | 16 (51.6) |
| White | 12 (38.7) |
| Hispanic/Latino or Black/African American | 3 (9.7) |
| First-generation student, n (%) | 11 (35.5) |
| International student, n (%) | 13 (41.9) |

Students contributed between 7 and 129 daily summaries (average 56 observations per participant). Across all observations, average nightly sleep duration was 423·52 minutes (SD 80·60), corresponding to approximately 7·1 hours per night. Average daily step count was 7,661 (SD 4,587). High emotional distress was reported on 436 of 1,724 person-days (25·3%). Summary statistics for wearable-derived and behavioral measures are presented in Table 2, and distributions of continuous variables are shown in Figure 1.

**Table 2:** Summary of Wearable-Derived and Behavioral Measures.

| Characteristic | Overall (n = 1,724) |
| --- | --- |
| Average breathing rate, mean (SD) | 15.31 (1.26) |
| Body temperature score, mean (SD) | 92.67 (11.85) |
| Deep sleep duration (min), mean (SD) | 87.56 (23.99) |
| REM sleep duration (min), mean (SD) | 98.02 (29.64) |
| Total sleep duration (min), mean (SD) | 423.52 (80.60) |
| Lowest heart rate (bpm), mean (SD) | 56.08 (6.99) |
| Recovery index score, mean (SD) | 72.71 (26.22) |
| Daily steps, mean (SD) | 7,661.36 (4,586.70) |
| Average HRV (ms), mean (SD) | 50.41 (19.45) |
| High emotional distress, n (%) | 436 (25.3) |

**Figure 1:**
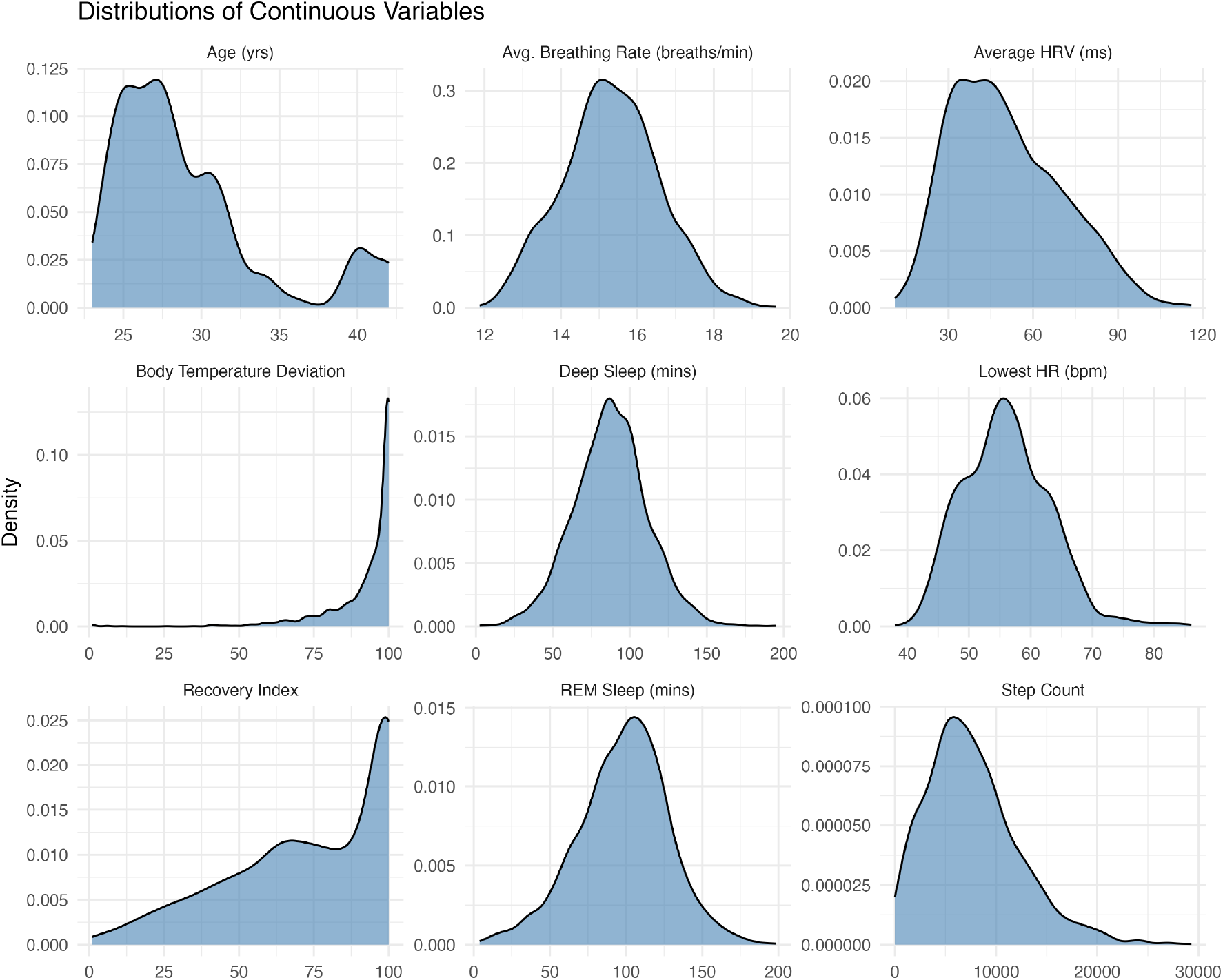
Distributions of continuous wearable-derived variables across all person-days.

Associations between covariates and log-transformed nightly HRV were examined at the 25th, 50th, and 75th percentiles (Table 3). This approach allowed assessment of whether associations differed across levels of physiological stress, with the lower quantile reflecting days characterized by relatively lower HRV (greater physiological stress). The outcome (HRV) is log-transformed to normalize the skewed distribution of HRV. Because of this transformation, the regression coefficients are interpreted as approximate percent changes in HRV associated with a one-unit increase in the covariate throughout the results.

**Table 3:** Quantile QIF estimates for log average HRV.

| Variable | $\tau = 0.25$ | $\tau = 0.50$ | $\tau = 0.75$ |
| --- | --- | --- | --- |
| Intercept | **3.840 (3.677, 4.003)** | **3.991 (3.936, 4.046)** | **4.193 (4.025, 4.361)** |
| Age | 0.005 (-0.173, 0.184) | **-0.039 (-0.050, -0.029)** | -0.060 (-0.138, 0.017) |
| Male | **-0.200 (-0.268, -0.133)** | **-0.175 (-0.236, -0.114)** | **-0.194 (-0.362, -0.026)** |
| International Student | -0.166 (-0.332, 0.001) | **-0.158 (-0.222, -0.094)** | -0.169 (-0.439, 0.101) |
| First-generation Student | -0.048 (-0.289, 0.193) | -0.023 (-0.047, 0.001) | -0.065 (-0.175, 0.046) |
| Average Breathing Rate | -0.004 (-0.033, 0.025) | **-0.018 (-0.021, -0.015)** | -0.011 (-0.032, 0.010) |
| Lowest Heart Rate | **-0.278 (-0.299, -0.258)** | **-0.275 (-0.282, -0.267)** | **-0.258 (-0.295, -0.220)** |
| Body Temperature | **0.028 (0.021, 0.034)** | **0.042 (0.039, 0.045)** | **0.019 (0.011, 0.027)** |
| Recovery Index | **0.024 (0.022, 0.026)** | **0.011 (0.009, 0.013)** | **0.026 (0.022, 0.030)** |
| Deep Sleep Duration | **0.026 (0.020, 0.033)** | **0.021 (0.014, 0.027)** | 0.003 (-0.008, 0.014) |
| REM Sleep Duration | **0.034 (0.025, 0.043)** | **0.026 (0.021, 0.030)** | **0.015 (0.008, 0.022)** |
| Daily Steps | 0.004 (-0.005, 0.012) | 0.002 (-0.002, 0.006) | **-0.019 (-0.026, -0.012)** |
| High Emotional Distress | **-0.094 (-0.111, -0.078)** | **-0.015 (-0.021, -0.008)** | **-0.022 (-0.030, -0.015)** |
| Steps × High ED | **0.058 (0.039, 0.077)** | **0.058 (0.050, 0.067)** | **0.074 (0.039, 0.108)** |

*Note.* Outcome is log-transformed nightly HRV. Values are regression coefficients with 95% confidence intervals in parentheses. Bold values indicate 95% confidence intervals that do not include 0.

Male students exhibited consistently lower HRV across quantiles compared with female students (25th: -22%, *β* = -0·200, CI: [-0·268, -0·133]; 50th: -19%, *β* = -0·175, CI: [-0·236, -0·114]; 75th: -21%, *β* = -0·194, CI: [-0·362, -0·026]), indicating a stable sex difference across the HRV distribution. At the median quantile, each additional year of age was associated with a 3·9% decrease in HRV (*β* = -0·039, CI: [-0·050, -0·029]), whereas age was not significantly associated with HRV in the lower quantile. International student status was associated with lower HRV at the median quantile (-17%, *β* = -0·158, CI: [-0·222, -0·094]), suggesting potential differences in physiological stress patterns by student status.

Sleep-related measures demonstrated consistent positive associations with HRV. Each additional minute of deep sleep was associated with a 2·6% increase in HRV in the lower quantile (*β* = 0·026, CI: [0·020, 0·033]) and a 2·1% increase at the median (*β* = 0·021, CI: [0·014, 0·027]). REM sleep duration was positively associated across quantiles (25th: +3·4%, *β* = 0·034, CI: [0·025, 0·043]; 50th: +2·6%, *β* = 0·026, CI: [0·021, 0·030]; 75th: +1·5%, *β* = 0·015, CI: [0·008, 0·022]), with larger magnitudes observed in the lower HRV quantile. Higher recovery index scores were similarly associated with higher HRV across quantiles. Lower nighttime heart rate demonstrated a strong inverse association with HRV throughout the distribution.

High emotional distress was inversely associated with HRV at all quantiles. The magnitude of this association was largest in the lower 25th HRV quantile (-9%, *β* = -0·094, CI: [-0·111, -0·078]), with smaller reductions observed at the median (-1·5%, *β* = -0·015, CI: [-0·021, -0·008]) and upper 75th quantiles (-2·2%, *β* = -0·022, CI: [-0·030, -0·015]).

A statistically significant interaction between daily step count and high emotional distress was observed across quantiles (25th: *β* = 0·058, 95% CI [0·039, 0·077]; 50th: *β* = 0·058, 95% CI [0·050, 0·067]; 75th: *β* = 0·074, 95% CI [0·039, 0·108]). Among low-to-moderate emotional distress days, physical activity was not positively associated with HRV and was negatively associated at the upper quantile (-1·9%, *β* = -0·019, 95% CI [-0·026, -0·012]). In contrast, on high emotional distress days, greater physical activity was associated with higher HRV (25th: +5·8%; 50th: +5·8%; 75th: +7·4%), with the largest effect observed in the upper HRV quantile.

On days of high emotional distress, greater physical activity was consistently associated with higher HRV, indicating a beneficial association under elevated emotional distress. The interaction pattern is illustrated in Figure 2.

**Figure 2:**
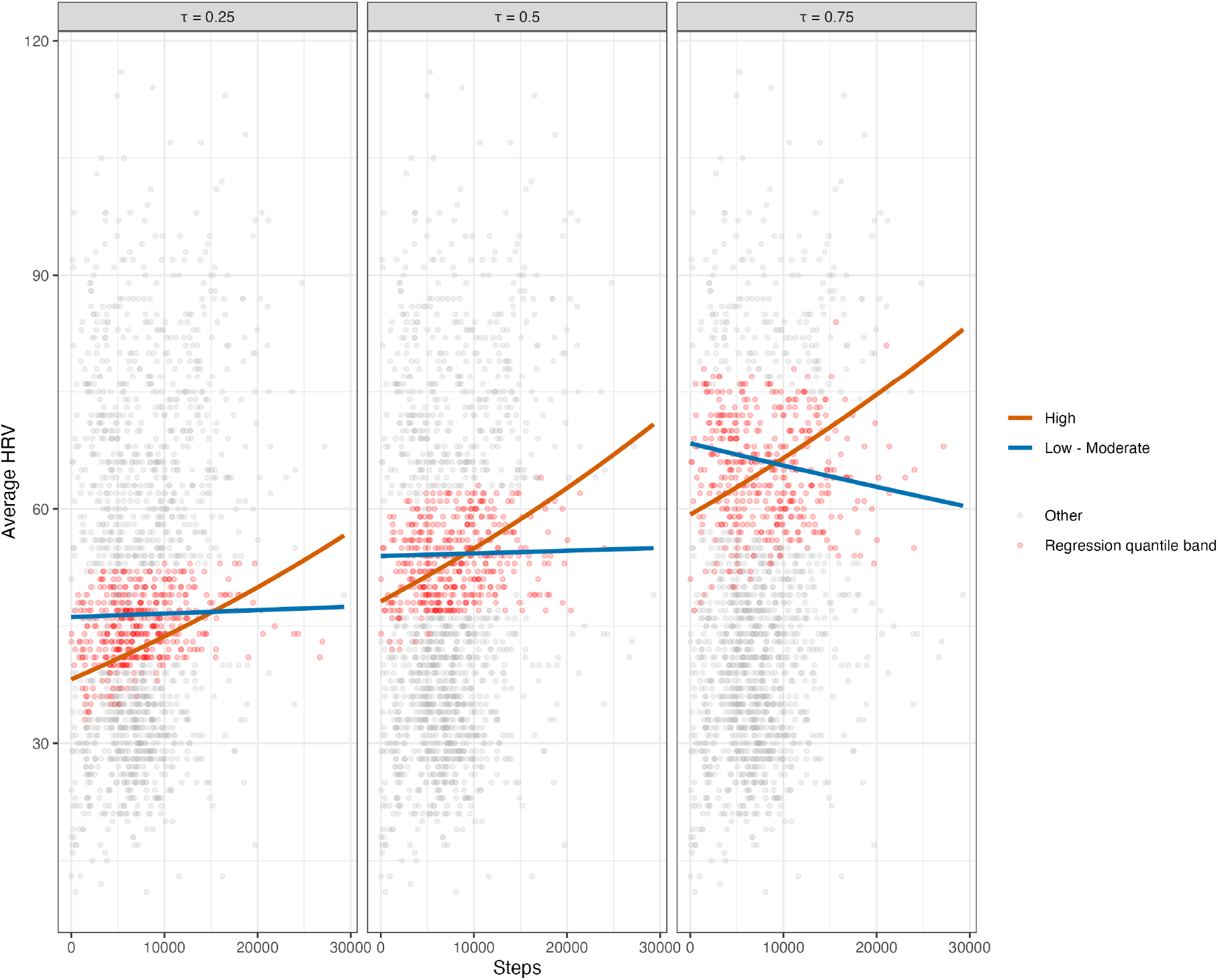
Estimated interaction between daily steps and emotional distress across quantiles of log average HRV. Red points indicate the 25% of observations lying closest to the fitted quantile regression line in each panel at *τ* = 0.25, 0.50, and 0.75.

## Discussion

In this prospective cohort study integrating wearable-derived physiological data with daily ecological momentary assessments, higher emotional distress was associated with lower nightly HRV, with the strongest effects observed on days characterized by already reduced HRV. These findings suggest that the physiological impact of emotional distress is amplified during periods of reduced autonomic regulatory capacity, highlighting vulnerable states in real-world settings.

While prior studies have documented inverse associations between psychological stress and HRV [14, 33], these relationships are typically evaluated at the mean level. By examining multiple quantiles of the HRV distribution, our analysis shows that the association between emotional distress and HRV varies across physiological states. The stronger inverse association at lower HRV quantiles suggests that stress-related autonomic dysregulation may compound when baseline regulatory capacity is already compromised.

A key finding is the context-dependent interaction between physical activity and emotional distress. Higher daily step counts were associated with higher HRV on days of elevated emotional distress, consistent with a buffering effect under acute stress. In contrast, this association was weaker or, in some cases, negatively associated on low-to-moderate distress days, particularly at higher HRV quantiles. This extends prior work showing beneficial effects of physical activity on HRV [17, 34] by demonstrating that these effects are most pronounced during periods of heightened emotional distress. Similar heterogeneity in physiological responses to behavioral factors has also been reported in wearable-based longitudinal studies [26, 35]. Overall, these results suggest that physical activity interventions may be most effective when timed to periods of elevated emotional distress.

Consistent with established literature, sleep-related measures, including deep sleep duration and recovery index, were positively associated with HRV across quantiles, reinforcing the role of sleep in autonomic regulation [35]. However, the observed positive association between REM sleep and HRV differs from some prior findings and should be interpreted cautiously. This discrepancy may reflect differences in study population or measurement approach and warrants further investigation.

Male and international students exhibited lower HRV at select quantiles. Although exploratory, these findings may reflect subgroup differences in physiological stress. Prior work has documented sex differences in HRV [13], while international students have been shown to experience elevated psychological stress in academic settings, which may contribute to altered physiological stress responses. These observations warrant further investigation in larger and more diverse samples.

Our findings support a digital phenotyping framework in which wearable-derived physiological signals and ecological momentary assessments can be used to identify high-risk stress states in real time. The observed buffering effect of physical activity on high emotional distress days suggests that wearable systems could trigger just-in-time adaptive interventions, such as physical activity prompts, during periods of elevated stress, supporting a precision mental health approach using passive, real-world physiological monitoring.

Limitations include the use of a convenience sample from a single public institution, which may limit generalizability. The small number of participants from underrepresented racial and ethnic groups limited our ability to conduct subgroup analyses, and collinearity between first-generation status and race/ethnicity further constrained interpretation.

In conclusion, emotional distress was inversely associated with nightly HRV, with stronger effects observed during periods of reduced autonomic functioning. Physical activity moderated this relationship under conditions of high emotional distress, supporting a context-dependent interaction. By leveraging wearable-derived physiological data and heterogeneous longitudinal modeling, this study advances understanding of stress processes in graduate students and highlights opportunities for adaptive, real-time interventions.

### Contributors

AQ and YG conceptualized and designed the study. CP led data collection, data management, and statistical analysis. AQ supervised statistical methodology and interpretation of results. CP drafted the initial manuscript. YG led on Ecological Momentary Assessment design, IRB approval and protocol for recruiting participants and critically revised the manuscript. CP and AQ accessed and verified the underlying data. All authors had full access to all the data in the study, contributed to data interpretation, reviewed and approved the final manuscript, and had final responsibility for the decision to submit for publication.

## Data Availability

The de-identified SMILES dataset, including demographic information and wearable-derived variables included in the analysis, will be made available upon reasonable request beginning at the time of publication. Data will be shared with qualified researchers for non-commercial research purposes following execution of a data use agreement. Requests should be directed to the corresponding author. The statistical analysis code is available upon request. The study protocol and analysis plan are available from the corresponding author on request. Individual participant data with identifiers will not be shared.

## Acknowledgments

This research was supported by National Cancer Institute SCH: Individualized Learning and Prediction for Heterogeneous Multimodal Data from Wearable Devices (R01: CA297869 to AQ and YG), National Science Foundation Data integration for heterogeneous data: A general framework for distribution shift, posterior drift and block missing data (DMS: 2515275 to AQ).

